# Spatial distribution and socioeconomic correlates of anaemia among women of reproductive age in Ghana: a hotspot analysis of the 2022 Demographic and Health Survey

**DOI:** 10.64898/2026.10.05.26364763

**Authors:** Abubakar Iddrisu Siddiq, Theophilus Arthur-Gyimah, Grace De-Graft Aitee, Oku Jeremiah Nii Annan Jeremiah, Emmanuel Vondee, Godlove Osei Asumang

## Abstract

**Background:** Anaemia in women of reproductive age is a persistent contributor to maternal morbidity and adverse birth outcomes in sub-Saharan Africa. Ghanaian surveys have repeatedly placed national anaemia prevalence in the World Health Organization’s severe public health problem range, yet the geographical distribution of this burden, and the socioeconomic factors that may account for it after geographical adjustment, remain insufficiently elucidated.

**Methods:** This cross-sectional study analysed the women’s and geospatial files of the 2022 Ghana Demographic and Health Survey. Anaemia was defined as altitude- and smoking-adjusted haemoglobin below World Health Organization thresholds among 7,557 women aged 15–49 years with a valid measurement. Survey-weighted multivariable logistic regression, accounting for clustering, stratification, and sampling weights, was used to elucidate socioeconomic and demographic correlates. Cluster-level prevalence was tested for spatial clustering with global Moran’s I and local Getis-Ord Gi* statistics (k-nearest neighbour weights, k = 8).

**Results:** Weighted anaemia prevalence was estimated at 41.1% (mild: 22.6%, moderate: 17.3%, and severe: 1.1%). Pregnant women appeared to have higher adjusted odds of anaemia than non-pregnant women (adjusted odds ratio [AOR] 1.71, 95% CI 1.38–2.12), as did women in the Northern (AOR 1.54, 95% CI 1.16–2.06) and Savannah (AOR 1.35, 95% CI 1.04–1.76) regions relative to Greater Accra, while the Ahafo region showed lower odds (AOR 0.64, 95% CI 0.43–0.94). Associations with rural residence, education, and wealth that were significant in crude analysis appeared substantially attenuated after adjustment. Moran’s I suggested significant positive spatial autocorrelation (I = 0.156, p = 0.001); Getis-Ord Gi* identified 109 hotspot and 117 cold spot clusters among 618 clusters, with hotspots concentrated in the north and cold spots in the mid-southern forest belt.

**Conclusion:** Two in five women of reproductive age in Ghana were estimated to have been anaemic in 2022. Risk appeared to be shaped more by region and pregnancy status than by household wealth or education once geography was accounted for. Anaemia-control programming may benefit from prioritising the identified northern hotspot areas alongside routine antenatal supplementation.

## Introduction

Anaemia, defined by the World Health Organization as a haemoglobin concentration below an age-, sex-, and physiology-specific threshold, remains among the most widespread nutritional and public health problems affecting women of reproductive age (1). It is not a single disease but the common endpoint of several overlapping causes, principally iron deficiency, malaria, other parasitic infections, chronic inflammation and haemoglobinopathies, and their relative contributions differ by setting (2). Globally, anaemia affects roughly three in ten women of reproductive age, with the burden concentrated disproportionately in low- and middle-income countries; sub-Saharan Africa carries one of the highest regional prevalences worldwide (3,4).

The consequences extend beyond a laboratory value. In pregnancy, anaemia is associated with an increased risk of low birth weight, preterm delivery and maternal mortality from haemorrhage, and in non-pregnant women, it is linked to fatigue, reduced physical work capacity and impaired cognitive performance, with adverse effects on household productivity and child care (5,6). Because women of reproductive age transition between pregnancy and non-pregnancy across the life course, population-level anaemia control may also constitute a form of preparedness for the physiological demands of pregnancy.

In Ghana, successive Demographic and Health Surveys have shown anaemia prevalence among women persisting in the range the World Health Organization classifies as a severe public health problem (7). The 2022 Ghana Demographic and Health Survey, whose data this paper analyses, reported anaemia in roughly two in five women overall, with a higher burden among pregnant women than non-pregnant women (8). This persistence, despite decades of national nutrition and malaria control programming, suggests that national averages alone do not adequately capture the factors sustaining anaemia in the Ghanaian population.

A substantial literature across Ghana and the wider sub-Saharan region has examined individual- and household-level correlates of anaemia in women, generally associating pregnancy status, rural residence, lower household wealth, lower educational attainment, and high parity (9,10). A smaller but growing body of work has moved beyond individual predictors to ask whether anaemia clusters geographically in ways that a conventional regression, which treats every woman as an independent observation, cannot show. Geographically weighted and hotspot analyses of Demographic and Health Survey data from Ethiopia and elsewhere in the region suggest that both the prevalence of anaemia and the strength of its socioeconomic correlates vary markedly across space, and that administrative units with the highest anaemia burden are not always the same units that conventional poverty targeting would prioritise (11,12). Within Ghana specifically, the north-south gradient in poverty, food insecurity, and child malnutrition is well documented (13), but no published study, to our knowledge, has formally tested for statistically significant spatial clusters, or hotspots, of anaemia among Ghanaian women of reproductive age using the geocoded 2022 survey round.

This evidence gap consequently warrants further investigation. Ghana’s national anaemia prevalence figure, however precisely measured, is a population average that can obscure sub-national concentrations of risk; a district or cluster with an average burden can sit beside one with a substantially higher burden without either being visible in the national statistic. Anaemia control resources, from iron-folic acid supplementation and malaria chemoprevention to nutrition education, are finite and are typically allocated at the regional or district level. Identifying whether Ghana’s anaemia burden is randomly scattered or forms statistically significant geographic clusters, and whether those clusters coincide with, or diverge from, conventional socioeconomic risk markers, may have a direct bearing on how such resources might reasonably be targeted.

This study therefore aimed to describe the prevalence and severity of anaemia among women aged 15–49 years in Ghana using the 2022 Demographic and Health Survey, to examine its socioeconomic and demographic correlates using survey-weighted multivariable logistic regression, and to test for and map statistically significant spatial clusters of anaemia prevalence at the survey-cluster level using Getis-Ord Gi* statistics. Specifically, the study sought to: (1) estimate the weighted prevalence and severity distribution of anaemia among women of reproductive age; (2) identify socioeconomic and demographic factors independently associated with anaemia; and (3) determine the spatial distribution of anaemia and identify statistically significant hotspot and cold spot clusters across the country.

## Methods

### Study design and setting

The present study comprised a secondary analysis of cross-sectional data from the 2022 Ghana Demographic and Health Survey (GDHS), a nationally representative household survey implemented by the Ghana Statistical Service in partnership with the Ghana Health Service and ICF, with technical and financial support from the DHS Program (8). Fieldwork covered all 16 administrative regions of Ghana (8). The survey used a stratified two-stage cluster design: enumeration areas (clusters) were selected as primary sampling units within urban/rural strata in each region, followed by systematic selection of households within each selected cluster. Full details of the sampling frame, questionnaire content, and fieldwork procedures are documented in the official survey report (8).

This analysis used two publicly available components of the survey: the women’s Individual Recode file (GHIR8CFL), which contains interview and biomarker data for women aged 15– 49 years, and the geospatial covariates file (GHGE8AFL), which provides displaced global positioning system (GPS) coordinates for each survey cluster.

### Study population

The target population consisted of women of reproductive age (15–49 years) who were usually resident in or had stayed the night before the survey in sampled households across Ghana. The source population for this analysis was the 15,014 women who completed the individual woman’s questionnaire in the 2022 GDHS. The study population for the primary analysis was defined as the subset of these women who resided in households randomly selected for haemoglobin testing and who had a valid haemoglobin measurement recorded. The unit of analysis for the descriptive and regression analyses was the individual woman; the unit of analysis for the spatial hotspot analysis was the survey cluster (primary sampling unit).

### Eligibility criteria

Inclusion criteria: women aged 15–49 years who completed the individual interview in the 2022 GDHS, resided in a household selected for the haemoglobin testing subsample, and consented to and received a capillary blood haemoglobin measurement with a recorded, valid result.

The study excluded women who were not selected for the haemoglobin-testing subsample and women who, although selected, had no haemoglobin measurement because they refused testing (n = 91), were absent at testing (n = 22), or had an unrecorded result (n = 6).

### Sample size

This was a secondary analysis; accordingly, no new sample size was calculated, and the analytic sample was determined by the original Ghana Demographic and Health Survey (GDHS) 2022 sampling design and haemoglobin-testing completeness. The Ghana Statistical Service and ICF determined the overall GDHS 2022 sample size using standard DHS Program sample size procedures (14). Of the 15,014 women interviewed, 7,676 resided in households randomly selected for haemoglobin testing; of these, 7,557 (98.4%) had a valid haemoglobin measurement and formed the analytic sample for the individual-level analyses. All 618 survey clusters had at least four women with a valid haemoglobin result (median 12, interquartile range 10–15) and were retained for the cluster-level spatial analysis.

### Sampling and recruitment

The GDHS 2022 used a stratified, two-stage probability sampling design (14). In the first stage, enumeration areas (clusters) were selected with probability proportional to size within each of the 32 sampling strata, which were formed by crossing the 16 regions with urban and rural residence. In the second stage, households were systematically selected from a household listing conducted in each selected cluster. Every eligible woman aged 15–49 years in selected households was interviewed. A subsample of households was additionally selected for anthropometric and biomarker measurement, including haemoglobin testing by trained fieldworkers using a battery-operated HemoCue photometer on a capillary blood sample, following standard DHS Program biomarker protocols. No independent participant recruitment was undertaken in this study; rather, the GDHS 2022 field teams completed all sampling and recruitment before data release.

### Study variables and operational definitions

Primary outcome. The primary outcome was anaemia status, derived from haemoglobin concentration adjusted for altitude and smoking status using the standard Demographic and Health Surveys (DHS) Program adjustment algorithm (15). Consistent with World Health Organization thresholds (1), women were classified by severity as not anaemic, mildly anaemic, moderately anaemic, or severely anaemic, using survey variable v457. For the regression analyses, anaemia was dichotomised as anaemic (mild, moderate, or severe) versus not anaemic, consistent with the binary primary-outcome specification agreed for this analysis; the four-level severity distribution is reported descriptively as a secondary outcome.

Independent and candidate variables. Candidate socioeconomic and demographic correlates were: place of residence (urban/rural); region (16 categories, Greater Accra as reference); age group in five-year bands; highest educational level attained (no education, primary, secondary, higher); household wealth quintile (the DHS Program’s composite wealth index, constructed from household asset ownership and housing characteristics via principal components analysis and grouped into national quintiles); marital status (never in union, currently in union, formerly in union); occupation, grouped from the detailed International Standard Classification of Occupations-based survey codes into not working, agriculture, manual or skilled labour, sales and services, and professional/clerical/managerial categories; parity (number of children ever born, grouped as 0, 1–2, 3–4, 5 or more); and current pregnancy status. Body mass index was examined descriptively but not entered into the regression models, because it may itself be altered by pregnancy (16) and could duplicate, rather than adjust for, the pregnancy-status pathway already modelled directly.

Potential confounders. Age, region, residence, wealth, education, marital status, occupation, parity, and pregnancy status were considered simultaneously in the multivariable model as mutually adjusting covariates rather than as a strict exposure-confounder hierarchy, consistent with the descriptive, correlate-identification aim of this analysis rather than a single a priori causal exposure.

### Data collection instrument

Data collection employed the standard DHS Program Women’s Questionnaire, administered face-to-face by trained interviewers in the respondent’s preferred language, and a Biomarker Questionnaire for anthropometric and haemoglobin measurement. Both instruments appear to be standardised, previously validated DHS Program tools that have been used across numerous countries and survey rounds (17); country-specific adaptations for the 2022 round (translation, local occupation, and region codes) are documented in the survey’s final report. This study used no additional, investigator-designed instrument.

### Pre-testing, validity and reliability

The Ghana Statistical Service and ICF conducted questionnaire pre-testing, interviewer training, translation, and content-validity review as part of the standard DHS Program survey implementation protocol prior to this study’s involvement. No new validity or reliability testing, including Cronbach’s alpha, appears to have been conducted for this secondary analysis, since no new composite scale was constructed; the wealth index used is the DHS Program’s existing, previously validated asset-based index (18).

### Data collection procedure

Trained Ghana Demographic and Health Survey (GDHS) 2022 field teams conducted household listing, interviews, and biomarker measurement under the supervision of the Ghana Statistical Service. The study’s involvement commenced after the authors obtained public-release de-identified data from the DHS Program; the authors undertook no original data collection, respondent contact, or field supervision.

### Data management and statistical analysis

All analyses were conducted in Python 3 using pandas for data management, the samplics package for design-based (survey-weighted) estimation, and libpysal and esda for spatial statistics. Women’s individual sample weights (v005, rescaled by dividing by 1,000,000) were applied throughout the individual-level analyses to restore national representativeness; primary sampling unit (v021) and sample strata (v023) were incorporated into all inferential analyses using Taylor-series linearisation to obtain design-based standard errors, consistent with DHS Program guidance for complex-survey analysis (19).

#### Descriptive statistics

Sociodemographic characteristics and anaemia prevalence were summarised as weighted percentages with unweighted sample sizes (Table 1).

**Table 1.** Sociodemographic characteristics and weighted anaemia prevalence among women aged 15–49 years, Ghana Demographic and Health Survey (DHS) 2022 (n = 7,557).

| Characteristic | Level | Sample size<br>(unweighted) | Weighted<br>percentage | Anaemia<br>prevalence,<br>weighted<br>percentage |
| --- | --- | --- | --- | --- |
| Place of residence |  |  |  |  |
|  | Urban | 3,708 | 57.2 | 39.4 |
|  | Rural | 3,849 | 42.8 | 43.4 |
| Region |  |  |  |  |
|  | Greater Accra | 508 | 16.3 | 38.8 |
|  | Ashanti | 576 | 19.4 | 37.5 |
|  | Ahafo | 432 | 3.9 | 30.1 |
|  | Bono | 369 | 2.5 | 36.3 |
|  | Bono East | 428 | 2.1 | 35.6 |
|  | Central | 514 | 11.7 | 44.4 |
|  | Eastern | 441 | 8.2 | 37.5 |
|  | North East | 524 | 2.2 | 43.2 |
|  | Northern | 444 | 2.5 | 51.8 |
|  | Oti | 487 | 2.6 | 46.3 |
|  | Savannah | 582 | 7.4 | 48.4 |
|  | Upper East | 470 | 1.8 | 45.0 |
|  | Upper West | 501 | 4.3 | 47.0 |
|  | Volta | 398 | 4.4 | 43.0 |
|  | Western | 409 | 6.4 | 45.9 |
|  | Western North | 474 | 4.3 | 40.3 |
| **Age group**<br>**(years)** |  |  |  |  |
|  | 15–19 | 1,424 | 17.8 | 43.8 |
|  | 20–24 | 1,328 | 17.7 | 38.6 |
|  | 25–29 | 1,194 | 15.8 | 40.1 |
|  | 30–34 | 1,137 | 15.1 | 38.2 |
|  | 35–39 | 1,016 | 13.8 | 39.7 |
|  | 40–44 | 837 | 11.3 | 46.2 |
|  | 45–49 | 621 | 8.6 | 43.4 |
| **Highest**<br>**educational**<br>**attainment** |  |  |  |  |
|  | No education | 1,676 | 15.9 | 45.4 |
|  | Primary | 1,144 | 14.0 | 42.6 |
|  | Secondary | 4,103 | 60.3 | 40.3 |
|  | Higher | 634 | 9.8 | 36.8 |
| **Wealth quintile** |  |  |  |  |
|  | Poorest | 1,839 | 16.0 | 46.6 |
|  | Poorer | 1,661 | 17.8 | 41.8 |
|  | Middle | 1,519 | 20.4 | 40.8 |
|  | Richer | 1,391 | 23.4 | 39.3 |
|  | Richest | 1,147 | 22.4 | 38.8 |
| **Marital status** |  |  |  |  |
|  | Never in union | 2,441 | 34.7 | 40.4 |
|  | Currently in union | 4,485 | 55.2 | 41.3 |
|  | Formerly in union | 631 | 10.1 | 42.8 |
| **Occupation** |  |  |  |  |
|  | Not working | 1,677 | 21.9 | 42.2 |
|  | Agriculture | 1,783 | 17.3 | 43.7 |
|  | Manual/skilled labour | 833 | 10.3 | 43.4 |
|  | Sales and services | 2,677 | 41.7 | 39.9 |
|  | Professional/clerical/managerial | 520 | 7.9 | 36.7 |
|  | Other/not classified | 67 | 1.0 | 37.1 |
| **Parity (children ever born)** |  |  |  |  |
|  | 0 | 2,311 | 32.0 | 41.3 |
|  | 1–2 | 2,189 | 29.7 | 39.3 |
|  | 3–4 | 1,695 | 22.4 | 39.9 |
|  | 5+ | 1,362 | 15.8 | 45.9 |
| **Current pregnancy status** |  |  |  |  |
|  | No/unsure | 7,004 | 93.3 | 40.4 |
|  | Yes | 553 | 6.7 | 51.4 |
| **Body mass index category (n = 7,550)** |  |  |  |  |
|  | Underweight | 577 | 7.0 | 49.3 |
|  | Normal | 4,151 | 49.4 | 44.6 |
|  | Overweight | 1,729 | 24.8 | 37.6 |
|  | Obese | 1,093 | 18.8 | 33.9 |

***BMI category is reported for 7,550 of 7,557 women (7 missing values).***

#### Bivariate analysis

The unadjusted (crude) association between each candidate variable and anaemia was estimated using single-variable survey-weighted logistic regression, yielding crude odds ratios (COR) with 95% confidence intervals and Wald p-values, rather than a design-naive chi-square test, so that the bivariate and multivariable estimates were derived under an identical, survey-consistent estimation framework (Table 2).

**Table 2.**
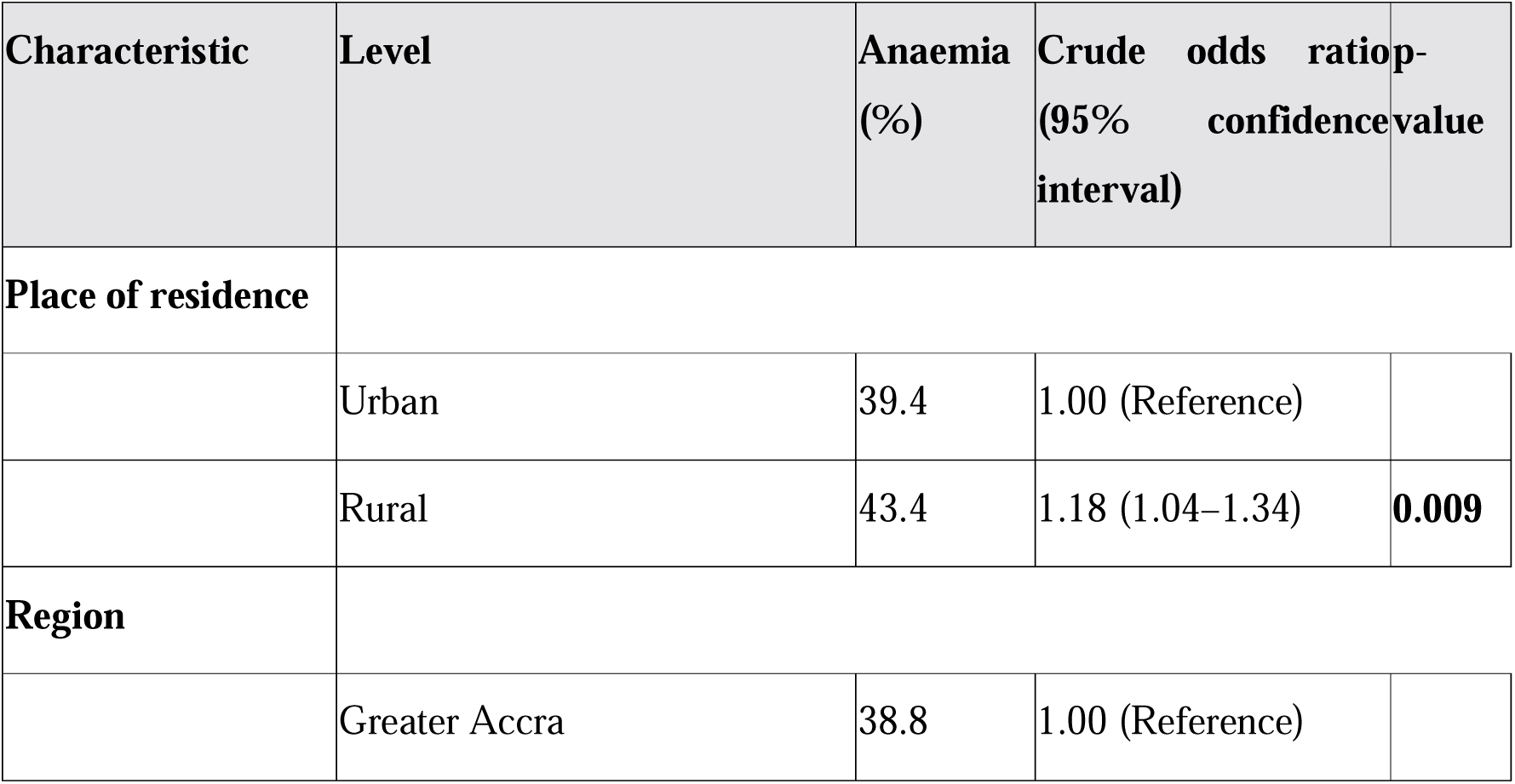
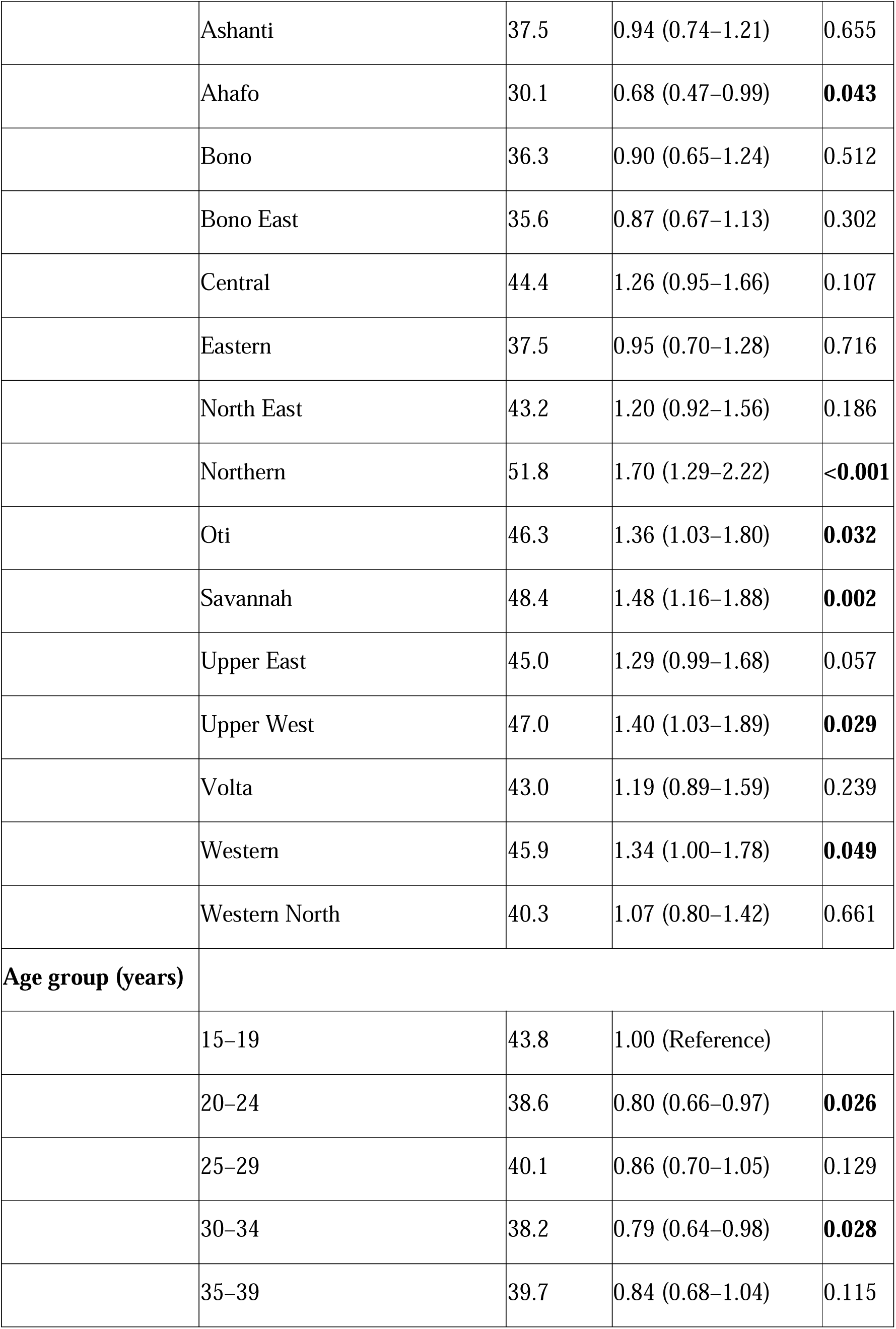
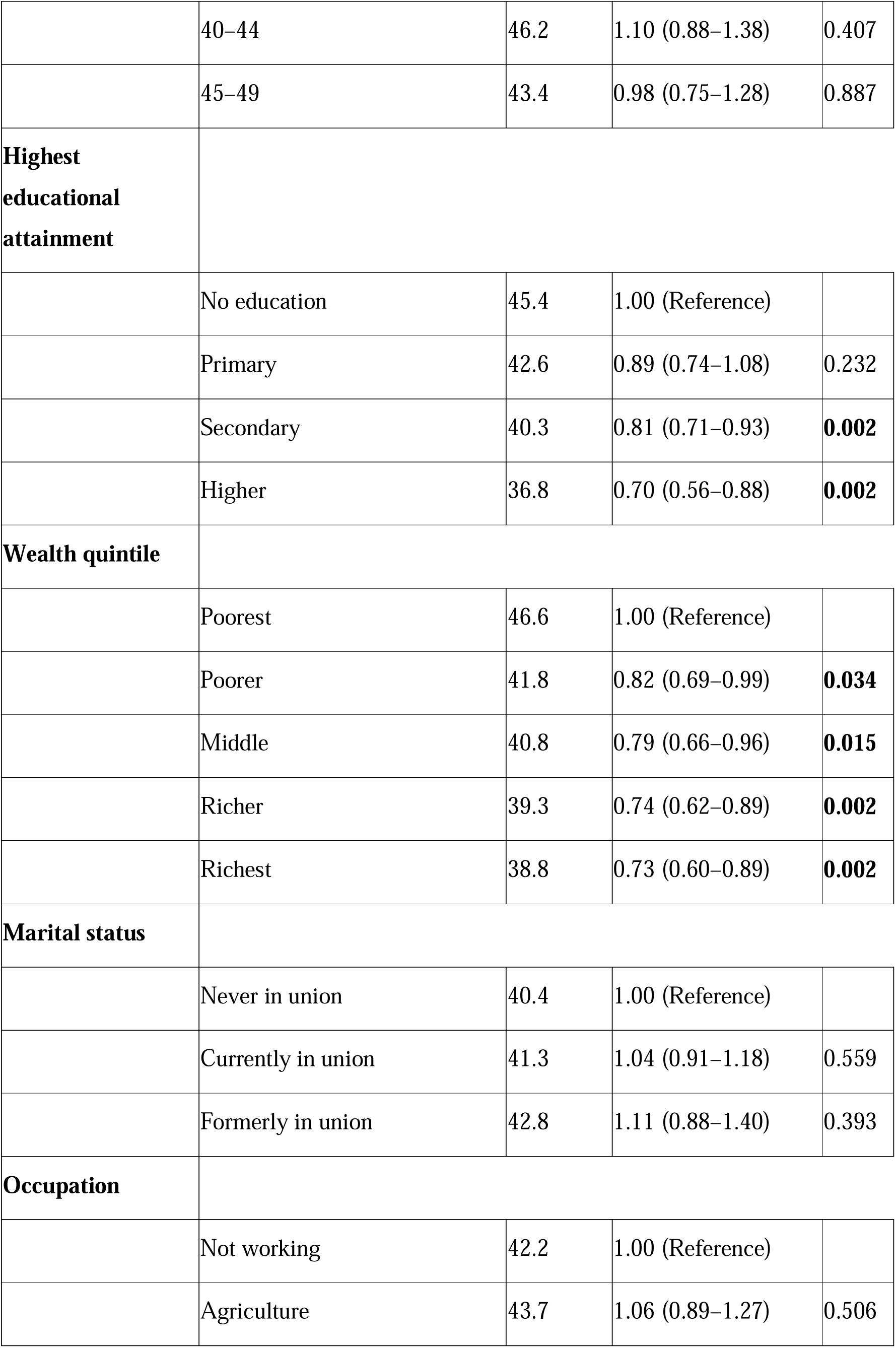
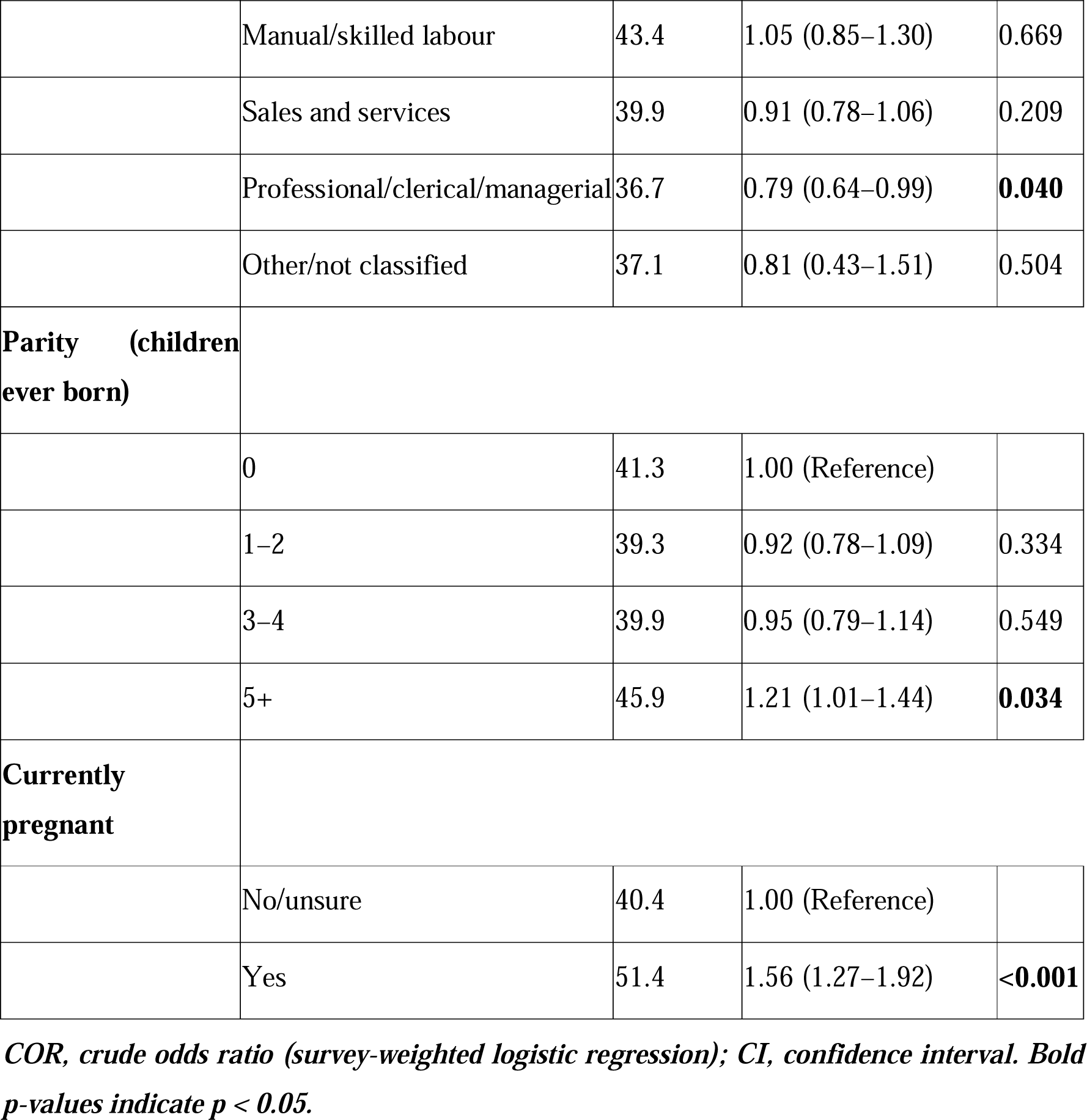
Bivariate (crude) analysis of factors associated with anaemia among women aged 15–49 years, Ghana DHS 2022.

#### Multivariable analysis

All candidate variables were entered simultaneously into a single survey-weighted multivariable logistic regression model to obtain adjusted odds ratios (AOR) with 95% confidence intervals (Table 2). Reference categories were selected as the largest or conceptually most neutral category of each variable (Table 2, footnote). Multicollinearity among candidate predictors was assessed using variance inflation factors (VIF) on the corresponding unweighted dummy-coded design matrix; all VIF values were below 6, which may suggest an absence of severe multicollinearity. Statistical significance was set at α = 0.05 throughout. Because the smallest observed p-values were below the precision of standard reporting, they are presented as p < 0.001 rather than p = 0.000.

#### Spatial analysis

Individual-level anaemia status was aggregated, using survey weights, to a weighted anaemia prevalence for each of the 618 survey clusters. These cluster-level prevalence estimates were merged with the corresponding displaced cluster GPS coordinates from the geospatial covariates file. Global spatial autocorrelation was assessed with Moran’s I; local spatial clustering was assessed with the Getis-Ord Gi* local statistic, using a k-nearest-neighbour spatial weights matrix with k = 8 neighbours per cluster. Statistical significance of the Gi* statistic for each cluster was assessed using a conditional permutation approach (999 permutations) and clusters were classified by confidence level (90%, 95%, 99%) as hotspots (significantly high prevalence surrounded by high-prevalence neighbours), cold spots (significantly low prevalence surrounded by low-prevalence neighbours), or not significant, following the standard Gi_Bin classification convention. Sensitivity of the hotspot classification to the choice of k was not formally assessed in this analysis.

### Bias and quality assurance

Several sources of potential bias were considered. Selection bias could arise if women selected for haemoglobin testing differed systematically from those excluded, or if non-response to testing (1.6% of the eligible sub-sample) was non-random; the GDHS 2022 biomarker sub-sample was itself randomly selected within the probability sampling design (20), and non-response was low, which may limit the scope for material selection bias. Measurement bias in haemoglobin readings was minimised through the use of a standardised, calibrated HemoCue device and trained fieldworkers following a uniform protocol across all clusters (20), though field conditions (temperature, device calibration drift) cannot be fully verified retrospectively from a secondary data source. Recall bias may affect self-reported variables such as parity and occupation, though these are relatively low-recall items compared with, for example, dietary intake. Social desirability bias is unlikely to materially affect the objective, device-measured primary outcome. Residual confounding from unmeasured factors, including malaria parasitaemia, dietary iron intake, helminth infection, and inflammatory markers, may nevertheless remain because these variables were unavailable in the files used for this analysis; this issue is addressed further under limitations.

### Ethical considerations

The present analysis was based on de-identified, publicly available secondary data. The original 2022 GDHS received ethical clearance from the Ghana Health Service Ethics Review Committee and from ICF’s Institutional Review Board in the United States, and all participants provided informed consent for interview and biomarker measurement at the time of the original survey (21). The DHS Program’s public-use data files contain no names, household addresses, or other direct identifiers; geographic identifiers are limited to the regional level in the survey record, and cluster GPS coordinates are randomly displaced (up to 2 km for urban clusters and up to 5 km, with 1% displaced up to 10 km, for rural clusters) so that individual enumeration areas cannot be re-identified (22). Access to the datasets used in this analysis was formally authorised by the DHS Program for the registered research project “Spatial distribution and socioeconomic correlates of anaemia among women of reproductive age in Ghana: a hotspot analysis using DHS 2022 geospatial data” (authorisation letter dated 3 September 2022, issued to Abubakar Iddrisu Siddiq, Presbyterian University, Ghana). No separate institutional ethical clearance appears to have been sought for this secondary analysis beyond DHS Program data-use authorisation.

## Results

### Participant flow

Of the 15,014 women aged 15–49 years interviewed in the 2022 GDHS, 7,676 resided in households selected for the haemoglobin testing sub-sample. Haemoglobin was not obtained for 119 of these women (91 refused, 22 were not present, and 6 for other reasons), leaving 7,557 women (98.4% of the eligible sub-sample) with a valid haemoglobin measurement, thereby constituting the analytic sample for the present study (Figure 1). All 618 survey clusters retained at least 4 tested women (median 12 women per cluster) and were used in the cluster-level spatial analysis.

**Figure 1.**
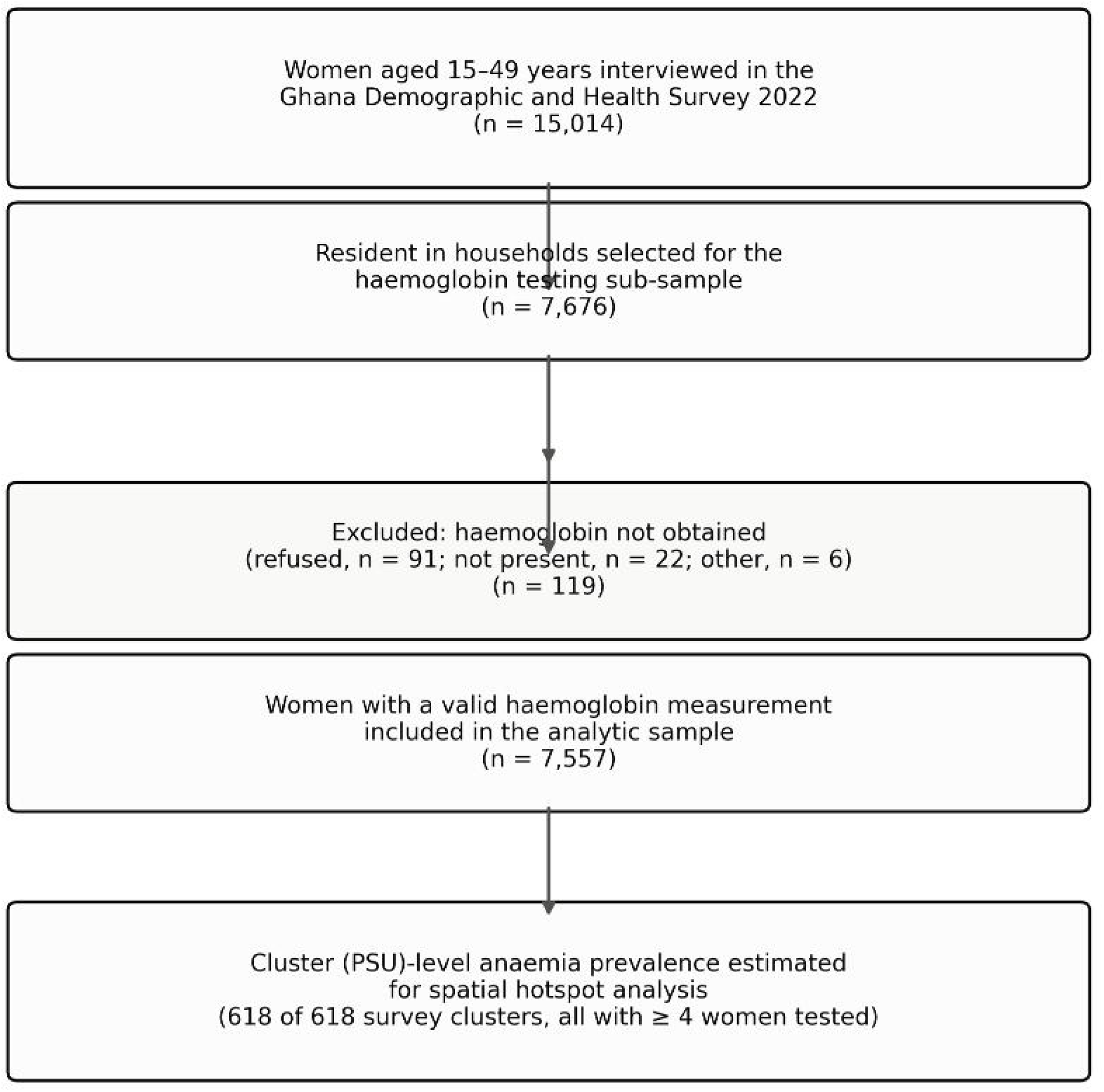
Flow diagram of participant inclusion in the analytic sample.

### Sociodemographic characteristics of study participants

Table 1 summarises the weighted sociodemographic characteristics of the analytic sample. Slightly more than half of women resided in urban areas (57.2%). The largest regional shares were from Ashanti (19.4%) and Greater Accra (16.3%); the smallest were from Bono East (2.1%) and Upper East (1.8%), reflecting the underlying regional population distribution captured by the survey’s sampling weights. Most women attained at least secondary education (60.3% secondary, 9.8% higher), while 15.9% had no formal education. Wealth quintiles were relatively evenly distributed, with a slightly larger share in the richer (23.4%) than the poorest (16.0%) quintile. Just over half of women (55.2%) were currently in a union, and 6.7% were pregnant at the time of interview.

### Prevalence and severity of anaemia

The weighted prevalence of anaemia among women aged 15–49 years was estimated at 41.1% (3,181 of 7,557 women; not adjusted for altitude and smoking, the corresponding unadjusted prevalence is not reported here as the DHS-standard adjusted classification was used throughout). The severity distribution suggests that 58.9% of women were not anaemic, 22.6% had mild anaemia, 17.3% had moderate anaemia, and 1.1% had severe anaemia (Figure 2). This computed prevalence appears closely consistent with the figure of approximately 41% reported in the official GDHS 2022 survey report (8) and with an independent secondary analysis of the same dataset (20), which similarly reported 40.4% among non-pregnant women and 51.4% among pregnant women; the corresponding figures in the present analysis were 40.4% and 51.4%, respectively, thereby providing a close internal consistency check on the survey-weighted computation used here.

**Figure 2.**
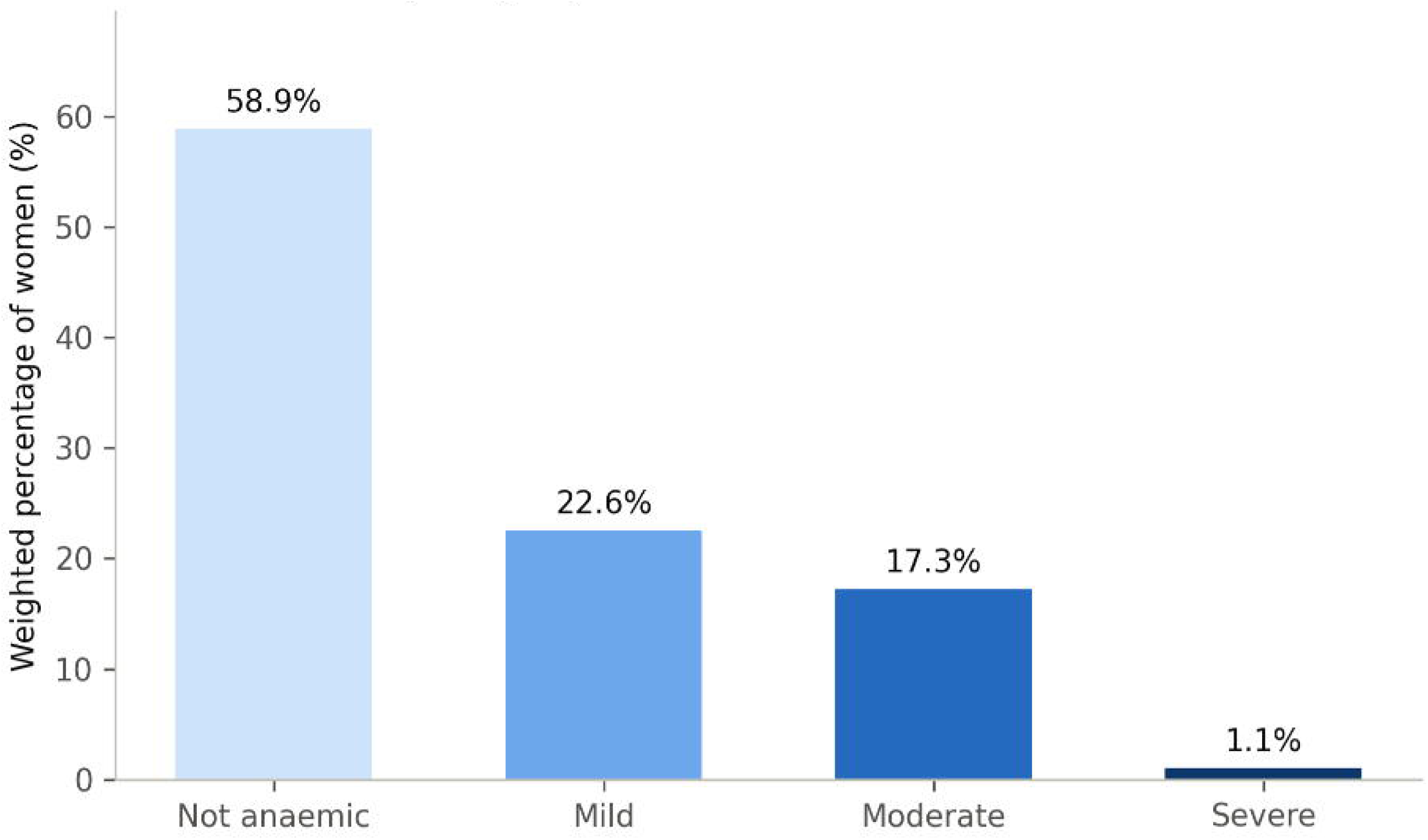
Severity distribution of anaemia among women aged 15–49 years, Ghana DHS 2022 (n = 7,557).

### Bivariate analysis of factors associated with anaemia

Table 2 delineates the weighted prevalence of anaemia and the crude (unadjusted) odds ratios for each candidate variable. In bivariate analysis, anaemia prevalence was significantly higher among rural women than among urban women (43.4% vs 39.4%; crude odds ratio [COR] 1.18, 95% confidence interval [CI] 1.04–1.34), among women with no education compared with those with higher education (45.4% vs 36.8%; COR for higher education 0.70, 95% CI 0.56–0.88), and among the poorest compared with the richest wealth quintile (46.6% vs 38.8%; COR for richest 0.73, 95% CI 0.60–0.89). Anaemia prevalence appeared to vary substantially by region, from 30.1% in Ahafo to 51.8% in the Northern Region. Pregnant women appeared to have higher unadjusted odds of anaemia than non-pregnant women (COR 1.56, 95% CI 1.27–1.92). Women with five or more children had higher odds than nulliparous women (COR 1.21, 95% CI 1.01–1.44).

### Multivariable analysis of factors associated with anaemia

In the multivariable model, adjusting simultaneously for residence, region, age, education, wealth, marital status, occupation, parity and pregnancy status, several associations observed in bivariate analysis appeared to be substantially attenuated (Table 2, Figure 3). Rural residence was no longer significantly associated with anaemia after adjustment (adjusted odds ratio [AOR] 1.07, 95% CI 0.91–1.26), nor were education (AOR for higher vs no education 0.95, 95% CI 0.67–1.35) or wealth quintile (AOR for richest vs poorest 0.87, 95% CI 0.66–1.16).

**Figure 3.**
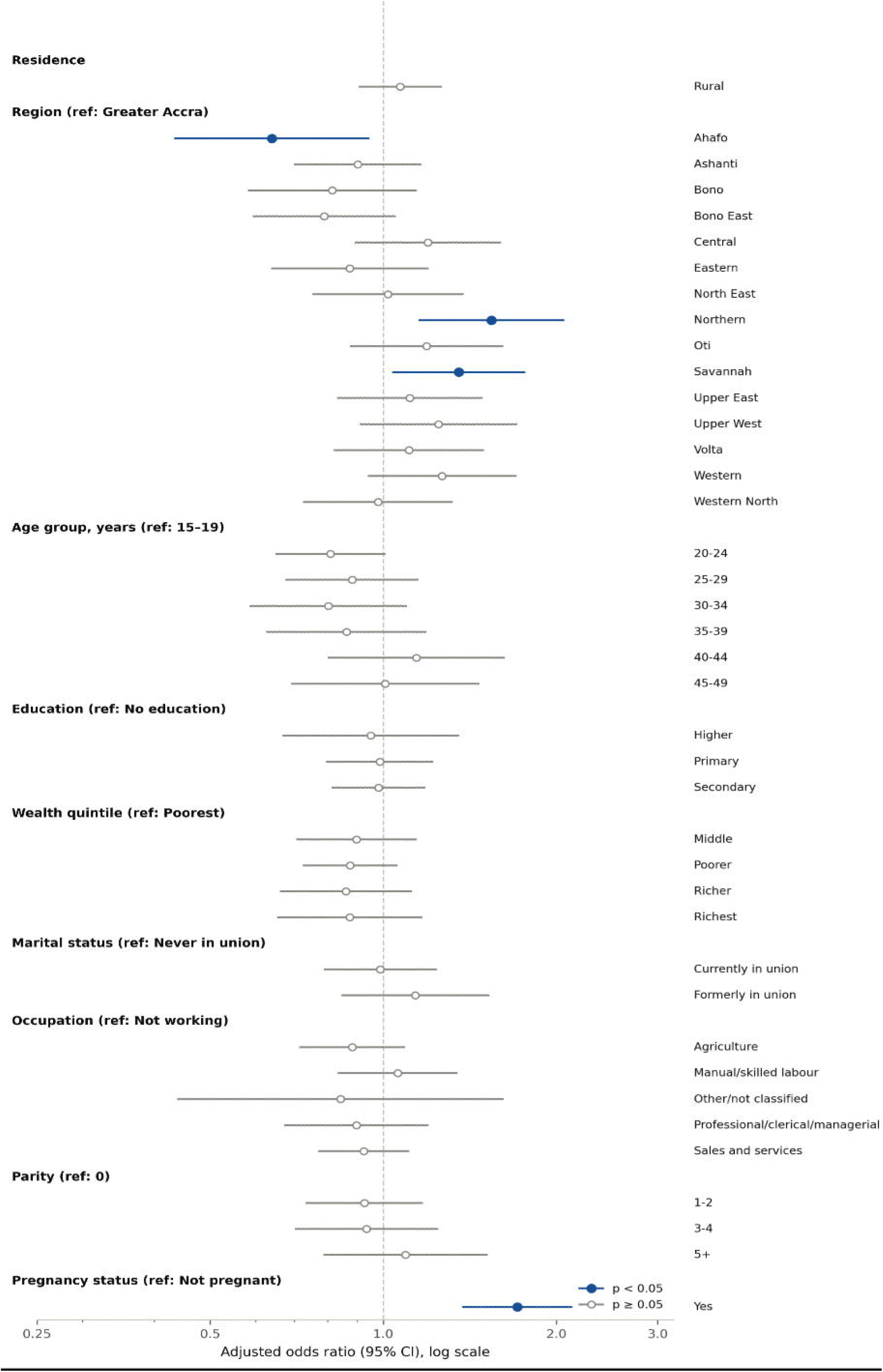
Adjusted odds ratios for socioeconomic and demographic correlates of anaemia among women aged 15–49 years, Ghana DHS 2022 (multivariable survey-weighted logistic regression).

Region appeared to remain independently associated with anaemia after adjustment. Relative to Greater Accra, women in the Northern region were estimated to have significantly higher adjusted odds of anaemia (AOR 1.54, 95% CI 1.16–2.06, p = 0.003), as did women in the Savannah region (AOR 1.35, 95% CI 1.04–1.76, p = 0.024). Conversely, women in the Ahafo region were estimated to have significantly lower adjusted odds than those in Greater Accra (AOR 0.64, 95% CI 0.43–0.94, p = 0.024). No other region differed significantly from Greater Accra after adjustment.

Pregnancy status appeared to exhibit the strongest independent association with anaemia in the multivariable model: pregnant women were estimated to have 71% higher adjusted odds of anaemia than non-pregnant women (AOR 1.71, 95% CI 1.38–2.12, p < 0.001). Age group, marital status, occupation and parity were not significantly associated with anaemia after adjustment for the other covariates (all p > 0.05; Table 2).

**Table 3.**
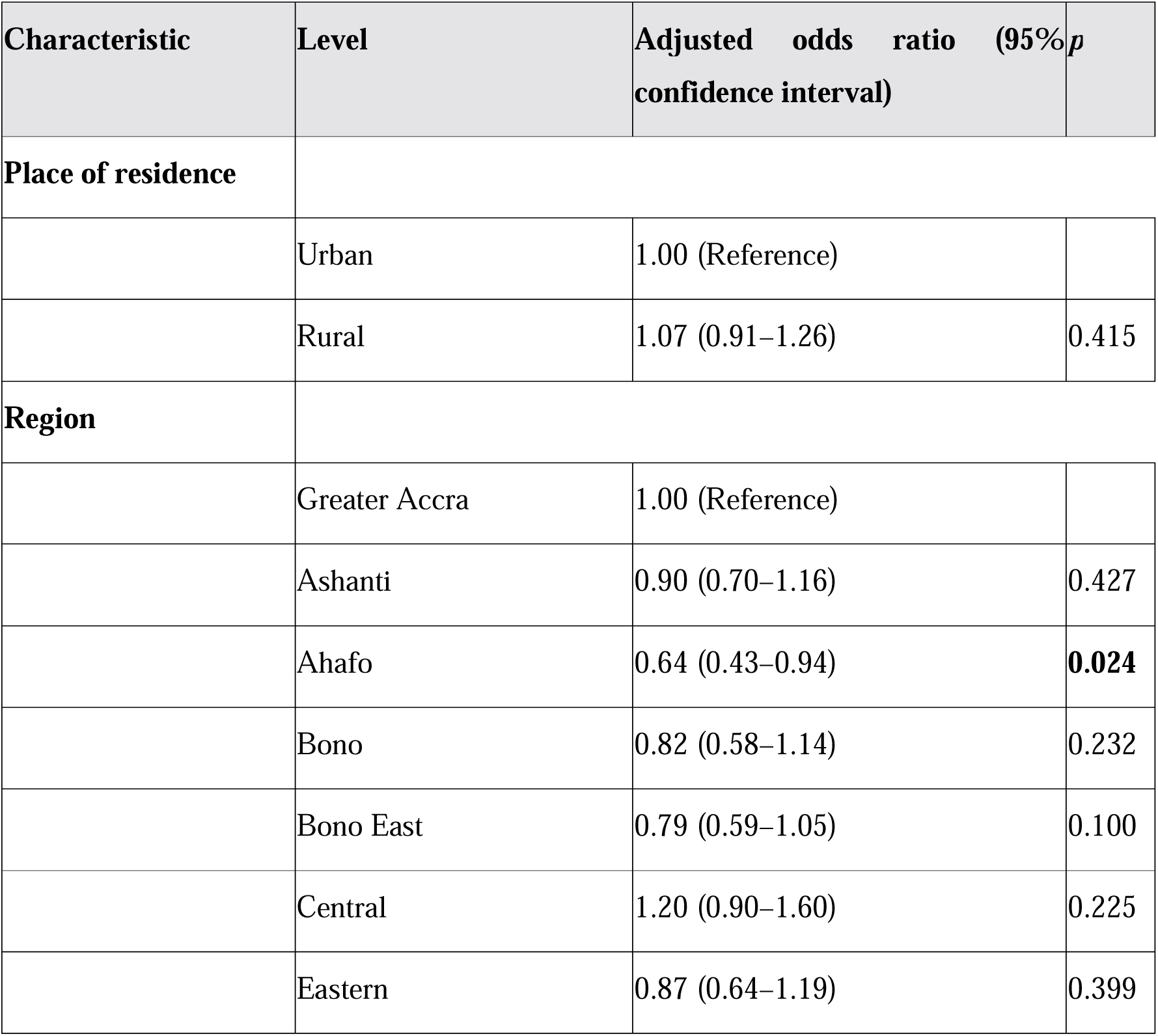
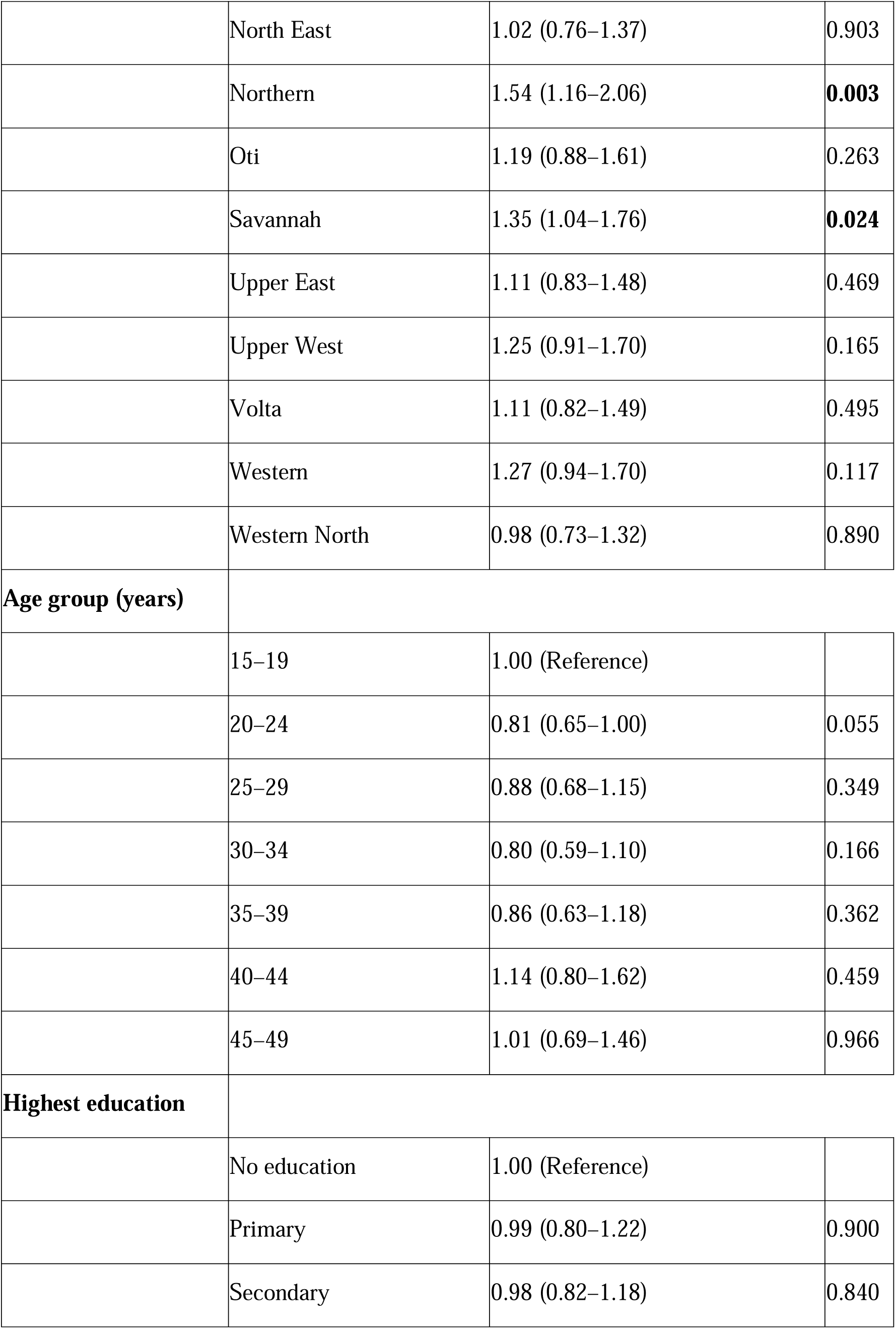
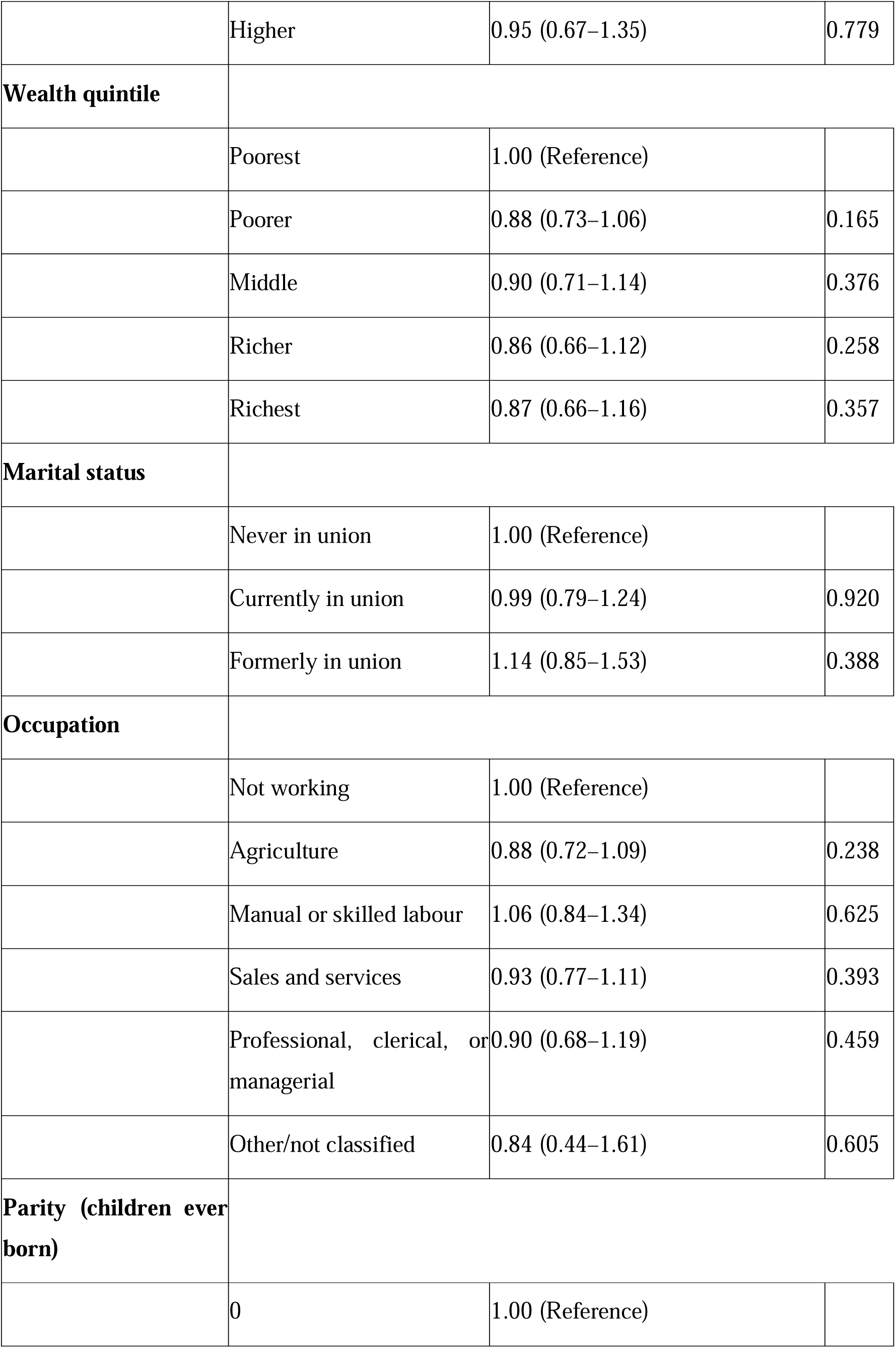
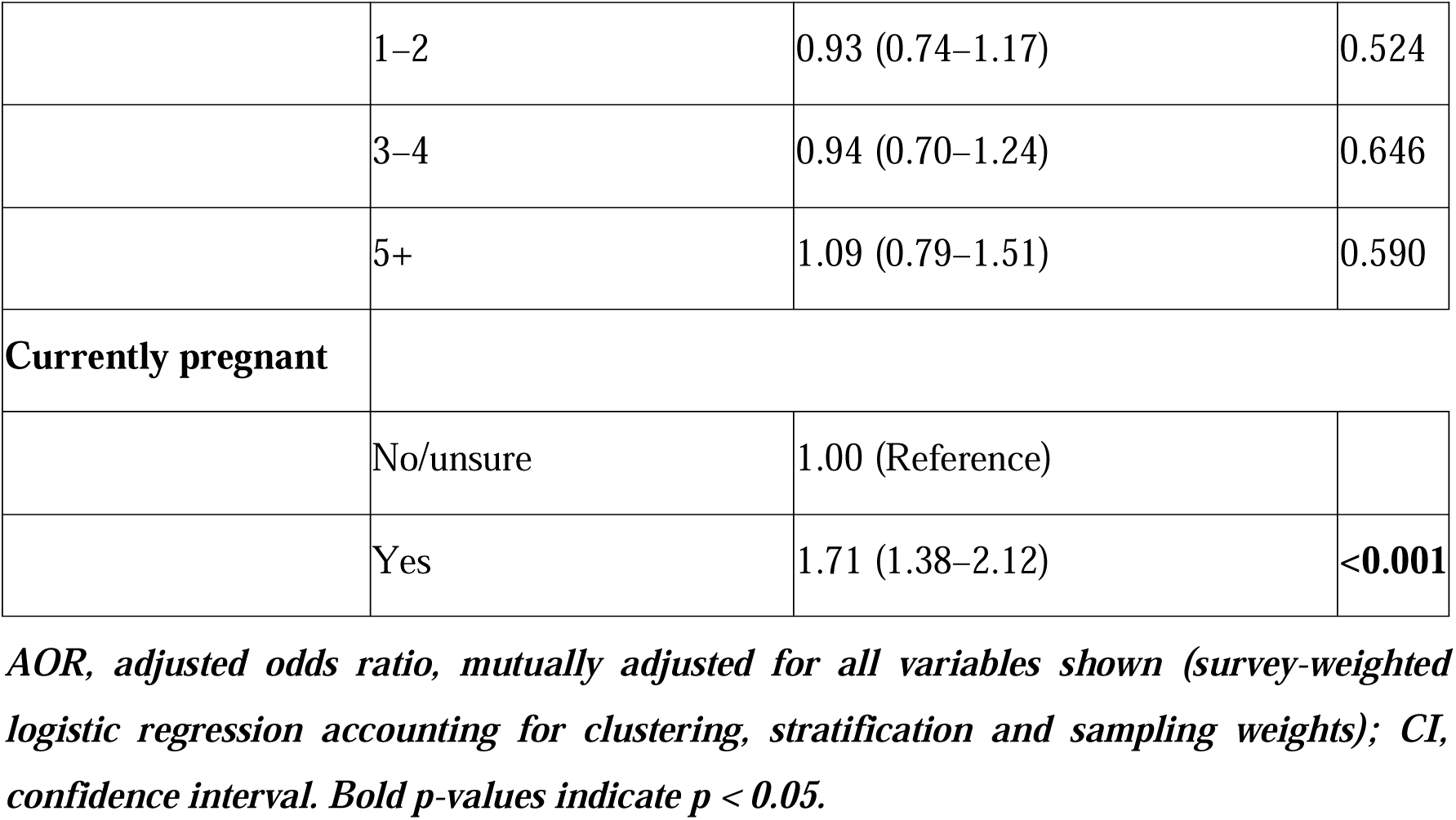
Multivariable (adjusted) analysis of factors associated with anaemia among women aged 15–49 years, Ghana DHS 2022.

| Characteristic | Level | Adjusted odds ratio (95% confidence interval) | $p$ |
| --- | --- | --- | --- |
| **Place of residence** |  |  |  |
|  | Urban | 1.00 (Reference) |  |
|  | Rural | 1.07 (0.91–1.26) | 0.415 |
| **Region** |  |  |  |
|  | Greater Accra | 1.00 (Reference) |  |
|  | Ashanti | 0.90 (0.70–1.16) | 0.427 |
|  | Ahafo | 0.64 (0.43–0.94) | **0.024** |
|  | Bono | 0.82 (0.58–1.14) | 0.232 |
|  | Bono East | 0.79 (0.59–1.05) | 0.100 |
|  | Central | 1.20 (0.90–1.60) | 0.225 |
|  | Eastern | 0.87 (0.64–1.19) | 0.399 |
|  | North East | 1.02 (0.76–1.37) | 0.903 |
|  | Northern | 1.54 (1.16–2.06) | **0.003** |
|  | Oti | 1.19 (0.88–1.61) | 0.263 |
|  | Savannah | 1.35 (1.04–1.76) | **0.024** |
|  | Upper East | 1.11 (0.83–1.48) | 0.469 |
|  | Upper West | 1.25 (0.91–1.70) | 0.165 |
|  | Volta | 1.11 (0.82–1.49) | 0.495 |
|  | Western | 1.27 (0.94–1.70) | 0.117 |
|  | Western North | 0.98 (0.73–1.32) | 0.890 |
| **Age group (years)** |  |  |  |
|  | 15–19 | 1.00 (Reference) |  |
|  | 20–24 | 0.81 (0.65–1.00) | 0.055 |
|  | 25–29 | 0.88 (0.68–1.15) | 0.349 |
|  | 30–34 | 0.80 (0.59–1.10) | 0.166 |
|  | 35–39 | 0.86 (0.63–1.18) | 0.362 |
|  | 40–44 | 1.14 (0.80–1.62) | 0.459 |
|  | 45–49 | 1.01 (0.69–1.46) | 0.966 |
| **Highest education** |  |  |  |
|  | No education | 1.00 (Reference) |  |
|  | Primary | 0.99 (0.80–1.22) | 0.900 |
|  | Secondary | 0.98 (0.82–1.18) | 0.840 |
|  | Higher | 0.95 (0.67–1.35) | 0.779 |
| **Wealth quintile** |  |  |  |
|  | Poorest | 1.00 (Reference) |  |
|  | Poorer | 0.88 (0.73–1.06) | 0.165 |
|  | Middle | 0.90 (0.71–1.14) | 0.376 |
|  | Richer | 0.86 (0.66–1.12) | 0.258 |
|  | Richest | 0.87 (0.66–1.16) | 0.357 |
| **Marital status** |  |  |  |
|  | Never in union | 1.00 (Reference) |  |
|  | Currently in union | 0.99 (0.79–1.24) | 0.920 |
|  | Formerly in union | 1.14 (0.85–1.53) | 0.388 |
| **Occupation** |  |  |  |
|  | Not working | 1.00 (Reference) |  |
|  | Agriculture | 0.88 (0.72–1.09) | 0.238 |
|  | Manual or skilled labour | 1.06 (0.84–1.34) | 0.625 |
|  | Sales and services | 0.93 (0.77–1.11) | 0.393 |
|  | Professional, clerical, or managerial | 0.90 (0.68–1.19) | 0.459 |
|  | Other/not classified | 0.84 (0.44–1.61) | 0.605 |
| **Parity (children ever born)** |  |  |  |
|  | 0 | 1.00 (Reference) |  |
|  | 1–2 | 0.93 (0.74–1.17) | 0.524 |
|  | 3–4 | 0.94 (0.70–1.24) | 0.646 |
|  | 5+ | 1.09 (0.79–1.51) | 0.590 |
| **Currently pregnant** |  |  |  |
|  | No/unsure | 1.00 (Reference) |  |
|  | Yes | 1.71 (1.38–2.12) | **<0.001** |

*AOR, adjusted odds ratio, mutually adjusted for all variables shown (survey-weighted logistic regression accounting for clustering, stratification and sampling weights); CI, confidence interval. Bold p-values indicate $p < 0.05$ .*

### Spatial distribution and hotspot analysis of anaemia

Cluster-level weighted anaemia prevalence ranged from 0% to 100% across the 618 survey clusters (median 40.0%). Global Moran’s I indicated statistically significant positive spatial autocorrelation in cluster-level anaemia prevalence (Moran’s I = 0.156, p = 0.001), suggesting that anaemia prevalence is not randomly distributed across Ghana but tends to cluster among geographically proximate survey locations.

The Getis-Ord Gi* local statistic identified 109 clusters (17.6% of all clusters) as statistically significant hotspots of high anaemia prevalence (23 at 99% confidence, 60 at 95% confidence, 26 at 90% confidence) and 117 clusters (18.9%) as statistically significant cold spots of low anaemia prevalence (35 at 99% confidence, 39 at 95% confidence, 43 at 90% confidence); the remaining 392 clusters (63.4%) showed no statistically significant local clustering (Figure 4). Hotspot clusters appeared to be concentrated in the northern regions, principally Northern (20 hotspot clusters), Western (15), North East (14), Oti (13) and Savannah (12) regions, broadly consistent with the elevated regional adjusted odds ratios described above, together with a secondary hotspot concentration observed in parts of the Western region. Cold spot clusters appeared to be concentrated in the mid-southern forest belt, principally Bono (22 cold spot clusters), Ahafo (16), Ashanti (14), Bono East (14) and Eastern (13) regions, together with Greater Accra (12). Hotspot clusters were split roughly evenly between rural (58) and urban (51) classifications, suggesting that the spatial clustering identified here reflects broader regional geography rather than the urban-rural distinction captured in the regression model.

**Figure 4.**
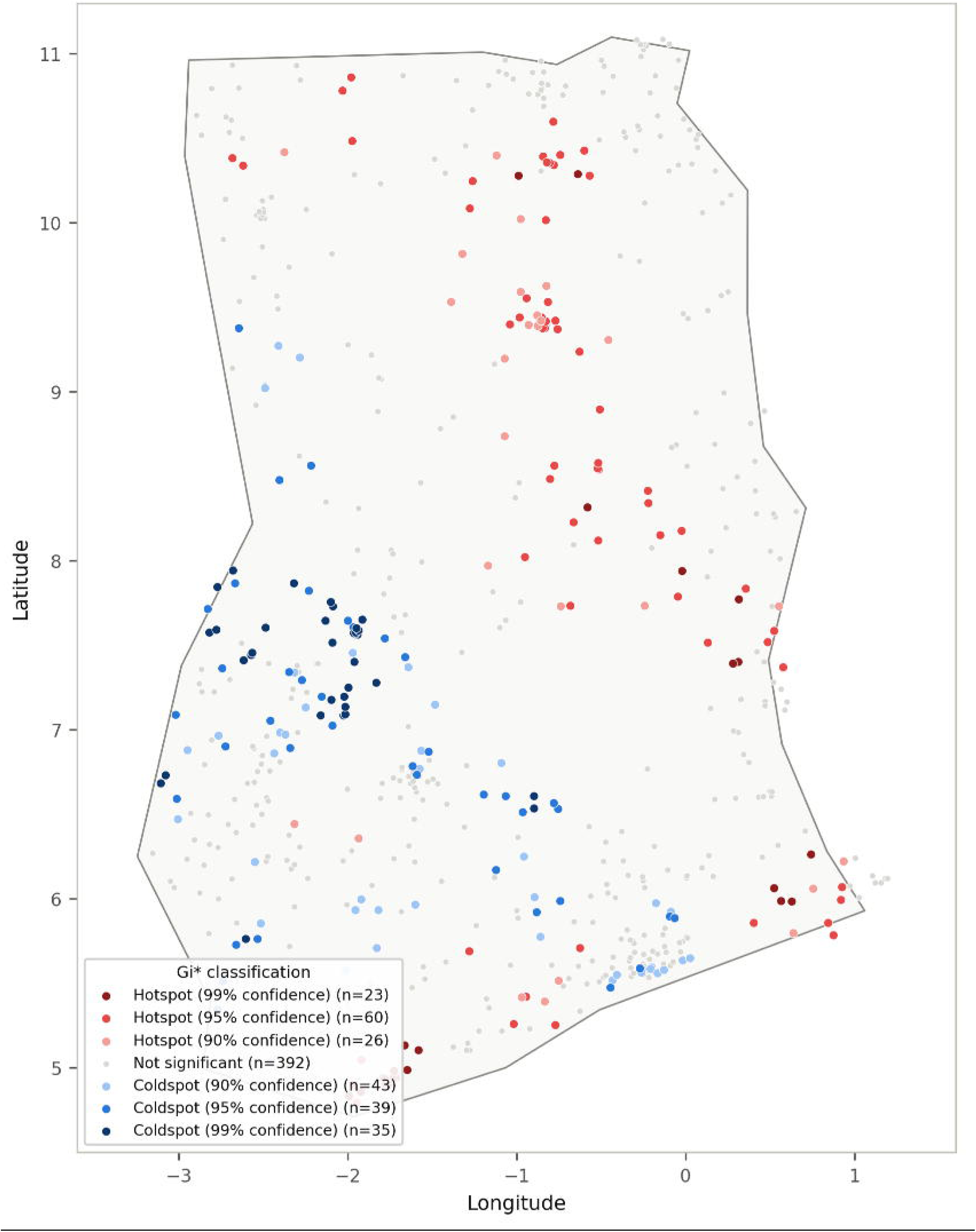
Getis-Ord Gi* hotspot analysis of cluster-level anaemia prevalence among women aged 15–49 years, Ghana DHS 2022 (k-nearest-neighbour spatial weights, k = 8).

## Discussion

This study found that two in five women of reproductive age in Ghana were anaemic in 2022, that this burden was shaped independently by pregnancy status and region rather than by individual household wealth or education after controlling for geography, and that anaemia appears to form statistically significant geographic hotspots concentrated in the country’s northern regions, with corresponding cold spots in the mid-southern forest belt.

The overall prevalence of 41.1% is consistent with the figure reported in the official GDHS 2022 report and an independent secondary analysis of the same survey round (8,20), which is unsurprising given that all three draw on the same underlying data, but it is a useful internal validity check on the survey-weighted estimation approach used here. This prevalence appears to remain within the World Health Organization’s severe public health problem category (≥40% prevalence) for the female population overall (23), and is considerably higher among pregnant women (51.4% in this analysis (20)). The persistence of this burden across successive DHS rounds, documented by (24) for Ghana specifically, suggests that national-level nutrition and malaria control programming, while likely beneficial, may not have been sufficient on its own to bring the national average below the severe threshold.

The finding that pregnancy status carried the strongest independent association with anaemia (AOR 1.71) is consistent with the physiological expectation that pregnancy increases iron and folate requirements while haemodilution lowers measured haemoglobin, and matches the pattern reported by (20) in their pregnancy-stratified analysis of the same survey. It supports the rationale for iron-folic acid supplementation throughout antenatal care as a first-line intervention, while also suggesting that a sizeable minority of non-pregnant women, four in ten in this sample, are anaemic outside of pregnancy and would not be reached by antenatal-only programming.

The attenuation of rural residence, education and wealth after adjusting for region is the most conceptually important finding of the multivariable analysis. In bivariate analysis, these variables showed the socioeconomic gradient reported in most of the sub-Saharan African literature (25), but each association weakened and lost statistical significance once region entered the model alongside the other covariates. Several strands of prior evidence help explain this pattern rather than treating it as anomalous. Multilevel analyses of pooled sub-Saharan African DHS data have found that a substantial share of the variance in anaemia risk sits at the community or cluster level, independent of individually measured household characteristics (26), which is consistent with region absorbing explanatory power that a model without a geographic term would otherwise attribute to wealth or education. Geographically weighted regression analyses in Ethiopia appear to indicate that the strength, and occasionally the direction, of the wealth-anaemia association is not constant across the country but varies by location (27), and a rural-urban stratified Ethiopian analysis similarly found that the wealth-anaemia relationship differs by residence stratum rather than operating as a single fixed effect (28). A coverage-equity analysis in Ethiopia likewise found that the areas of highest anaemia burden did not always coincide with the areas conventional poverty targeting would prioritise (29). Read together, this literature suggests that in agrarian, food-insecurity-driven settings such as northern Ghana, regional exposures operating largely independently of individual household wealth—plausibly seasonal food availability, malaria endemicity, dietary iron bioavailability, and distance to health facilities—may play a more significant role in determining anaemia risk than a household’s position in the national wealth distribution. This is a plausible interpretation rather than a demonstrated causal mechanism; distinguishing compositional effects (who lives where) from genuinely place-based effects would require the decomposition or multilevel modelling approaches used elsewhere in the literature (30), which were beyond the scope of this analysis.

The elevated adjusted odds of anaemia in the Northern and Savannah Regions, and the significant hotspot clusters detected there by the Getis-Ord Gi* analysis, echo a longstanding pattern in Ghana’s health and nutrition statistics. The north-south gradient in child malnutrition and poverty documented by (31) nearly two decades ago, and the persistent regional disparity in anaemia burden reported across multiple DHS rounds by (24), both point to a structural, rather than transient, feature of northern Ghana’s health landscape. The lower adjusted odds observed in the Ahafo Region, one of Ghana’s more recently created regions situated in the cocoa-growing forest belt, appear consistent with the cold spot concentration detected there and in neighbouring Bono, Bono East, and Ashanti Regions, an area generally associated with more diversified agriculture and food availability than the savannah north.

To our knowledge, this is among the first studies to formally test for, and map, statistically significant Getis-Ord Gi* hotspots of anaemia among women of reproductive age using the geocoded 2022 GDHS round specifically. The significant global Moran’s I (0.156, p = 0.001) suggests that this clustering is unlikely to be attributable to chance, and is directionally consistent with hotspot analyses of anaemia and related outcomes conducted elsewhere in the region using comparable DHS geospatial data (12). The roughly even urban-rural split among hotspot clusters (58 rural, 51 urban) is a notable finding in its own right: it suggests that the spatial clustering detected here reflects a broader regional geography of risk rather than simply redescribing the urban-rural divide already captured by the regression model, where it was found to be non-significant after adjustment.

### Strengths and limitations

This study has several strengths. It used a large, nationally representative sample with objectively measured haemoglobin rather than self-reported symptoms, applied a design-consistent survey-weighted estimation framework throughout both the regression and descriptive analyses, and combined individual-level regression with cluster-level spatial statistics to characterise the anaemia burden from two complementary angles. Using the GDHS 2022 geospatial file enabled the addition of a previously absent Ghana-specific spatial hotspot perspective on anaemia to an already substantial literature on individual-level correlates.

Several limitations should be considered when interpreting these findings. First, this is a cross-sectional study; all associations, including the regional and pregnancy-status associations, describe statistical relationships at a single point in time and cannot establish temporal sequence or causation. Second, several biologically plausible confounders and mediators of anaemia, including malaria parasitaemia, helminth infection, inflammatory markers, dietary iron and folate intake, and haemoglobinopathy status, were unavailable in the analytic dataset and could not be adjusted for; residual confounding from these factors, several of which plausibly vary by region, cannot be ruled out and may itself partly explain the regional associations observed. Third, cluster GPS coordinates in DHS Program data are randomly displaced by up to 2 km in urban clusters and up to 5 km (with 1% displaced up to 10 km) in rural clusters, to protect respondent confidentiality (22); this displacement may introduce a small amount of positional noise into the hotspot analysis, which is unlikely to materially affect regional-scale patterns but could attenuate very fine-grained local clustering. Fourth, haemoglobin was measured in a subsample of households rather than the full sample, and while non-response within this subsample was low (1.6%), any systematic difference between tested and untested women that is not captured by the survey weights could introduce a small selection bias. Fifth, the k = 8 nearest-neighbour specification used for the spatial weights matrix is one of several plausible choices, and the hotspot classification was not formally tested for sensitivity to this choice. Finally, occupation and parity categories with very small cell sizes (for example, the 67 women classified as having an unclassifiable occupation) yield wide confidence intervals for those specific categories, and results for such categories should be interpreted with caution.

### Implications for practice and policy

For antenatal and reproductive health services, the apparent independent association between pregnancy and anaemia reinforces existing guidance on universal iron-folic acid supplementation and early antenatal haemoglobin screening, while the substantial anaemia burden among non-pregnant women (40.4% in this sample) indicates that antenatal-only programming could leave many anaemic women of reproductive age unreached; preconception and general reproductive health platforms may be a relevant complementary channel.

For the Ghana Health Service and regional health directorates, the concentration of statistically significant anaemia hotspots in the Northern and North East regions (32) may provide an evidence base for considering differentiated, geography-informed resource allocation, for example, prioritising community-level iron supplementation, malaria chemoprevention and nutrition education outreach in identified hotspot districts, alongside the routine, universal components of anaemia control programming.

For the Ministry of Health and national nutrition policy more broadly, the finding that conventional household socioeconomic markers lost explanatory power once region was accounted for suggests that anaemia-control targeting based solely on household wealth proxies may miss a geographically concentrated component of the burden; complementing wealth-based targeting with the small-area spatial surveillance demonstrated here could improve the precision of programme targeting. These implications are proportionate to a single cross-sectional national survey round and should be read as evidence to inform, rather than substitute for, local programmatic and budgetary decision-making.

### Recommendations for future research

The limitations above suggest three directions for future research: First, linking haemoglobin and geospatial DHS data with concurrent malaria parasitaemia, inflammation and micronutrient biomarker data, where available in a given survey round, would allow the relative contribution of infectious and nutritional pathways to the observed regional clustering to be disentangled, rather than inferred indirectly as in this analysis. Second, a multilevel or spatial decomposition analysis, explicitly partitioning anaemia risk into individual-, cluster- and region-level components, would formally test whether the attenuation of socioeconomic associations observed here reflects genuine place-based effects or the compositional sorting of poorer, less-educated women into higher-risk regions, a distinction this cross-sectional regression could not make. Third, repeating this hotspot analysis using earlier GDHS rounds with geospatial data available would establish whether the identified hotspot and cold spot locations are stable over time, which would strengthen the case for using them to guide medium-term programme planning rather than reacting to what could otherwise be a single survey round’s sampling variability.

### Conclusion

Anaemia affected two in five women of reproductive age in Ghana in 2022, with pregnant women and women resident in the Northern and Savannah regions bearing a disproportionate share of that burden. Household socioeconomic markers that were associated with anaemia in unadjusted comparisons were not independently significant once region was accounted for, indicating that geography, rather than household wealth or education alone, may constitute a more consistent population-level marker of risk in this dataset. Statistically significant hotspots of high anaemia prevalence, concentrated in the country’s northern regions, and cold spots concentrated in the mid-southern forest belt, were identified using Getis-Ord Gi* analysis of the 2022 survey’s geocoded clusters. These findings may support continued prioritisation of antenatal iron-folic acid supplementation for pregnant women, and suggest that anaemia-control programming in Ghana may benefit from incorporating small-area spatial evidence alongside conventional household-wealth-based targeting, particularly in the identified northern hotspot regions. Based on a single cross-sectional survey round, this study describes an association pattern rather than a causal one, and its findings should be read as one input among several into programme and policy decisions.

## Data Availability

The datasets analysed in this study are publicly available from the Demographic and Health Surveys (DHS) Program (https://www.dhsprogram.com) upon registration and approval of a research project, subject to the DHS Program's data use terms (https://dhsprogram.com/Data/terms-of-use.cfm). The specific files used were the Ghana 2022 Individual Recode (GHIR8CDT) and geospatial covariates (GHGE8AFL) datasets. The redistribution of DHS microdata is restricted; interested researchers should apply directly to the DHS Program. Derived, non-identifying analysis code and aggregated cluster-level results are available from the corresponding author on reasonable request.

https://www.dhsprogram.com/data/dataset_admin/login_main.cfm

## Abbreviations

AOR: adjusted odds ratio
CI: confidence interval
COR: crude odds ratio
DHS: Demographic and Health Survey
GDHS: Ghana Demographic and Health Survey
Gi*: Getis-Ord Gi* local spatial statistic
GPS: Global Positioning System
GSS: Ghana Statistical Service
ICF: ICF International (implementing partner of the DHS Program)
PSU: primary sampling unit
STROBE: Strengthening the Reporting of Observational Studies in Epidemiology
VIF: variance inflation factor
WHO: World Health Organization

## Declarations

### Ethics approval and consent to participate

See Ethical considerations in the Methods section. This study used de-identified, publicly available secondary data released under DHS Program data-use authorisation; the original survey received ethical clearance from the Ghana Health Service Ethics Review Committee and ICF’s Institutional Review Board, and all original participants provided informed consent.

### Consent for publication

Not applicable (no data, images, or case details concerning identifiable individuals are presented).

### Authors’ contributions

Abubakar Iddrisu Siddiq led the conceptualization, methodology, statistical analysis, and preparation of the original draft; Theophilus Arthur-Gyimah contributed to data curation and validation; Grace De-Graft Aitee contributed to the literature review and interpretation of findings; Oku Jeremiah Nii Annan Jeremiah contributed to data visualization and results interpretation; Emmanuel Vondee contributed to methodological review and critical revision of the manuscript; and Godlove Osei Asumang contributed to supervision, project administration, and final manuscript review, with all six authors approving the final version.

